# Evaluating the effects of an intervention to improve the health environment for mothers and children in health centres (BECEYA) in Mali: a qualitative study

**DOI:** 10.1101/2022.04.04.22273417

**Authors:** Patrice Ngangue, Katherine Robert, Birama Apho Ly, Fatoumata Traoré, Maude Vezina

## Abstract

**Background:** An intervention aiming to improve maternal and children environment in healthcare facilities (BECEYA) was launched in three regions of Mali. This study aimed to explore the perceptions and experiences of patients and their companions, community actors and healthcare facilities staff on the effects of the BECEYA intervention in two regions of Mali.

**Methods:** We conducted a qualitative study using an empirical phenomenological approach. Through purposive sampling, women who attended antenatal care in the selected healthcare centres, companions, and health facility staff members were recruited. Data were collected during January and February 2020 through semi-structured individual interviews and focus groups. According to Braun & Clarke approach, audio recordings were transcribed verbatim, and a thematic analysis was conducted in five main steps.

**Results:** We recruited 26 participants in individual interviews and 20 participants in focus groups. Donabedian conceptual framework of quality of care was used to present the perceived changes following the implementation of the BECEYA project. The themes that emerged from data analysis are perceived changes in terms of infrastructure (perceived changes in the characteristics of the healthcare facilities setting, including the infrastructure introduced by the BECEYA project), process (changes in the delivery and use of care introduced or resulting from BECEYA activities), and outcome (the direct and indirect effects of these changes on the health status of patients and the population).

**Conclusion:** The study identified the positive effects of women users of the services, their companions and health centre staff following the implementation of the project. Therefore, this study contributes to the advancement of knowledge to show the link between improving the health environment of health centres in developing countries, a major aspect of the quality of care, and maternal and child health care.

## Background

Safe and adequate environmental conditions in health care facilities (HCFs) – including the availability of water, sanitation, hygiene (WASH), energy, and waste management are essential to protect and improve the health of patients, staff, visitors, and the wider community ^1^. Furthermore, ensuring access to water, sanitation and hygiene in health facilities is part of the United Nations agenda for global transformation through the Sustainable Development Goals (SDGs) ^2^. In addition, essential WASH services in health facilities and maternity wards are fundamental to providing quality care and ensuring the fulfilment of primary health care commitments ^3, 4^.

In healthcare facilities, WASH interventions aim to promote a healthy environment to minimise disease risk for users (patients, health care staff and maintenance staff) and the surrounding community. Interventions consist of providing a water point and improving the sanitary environment (including sanitation, availability of hand hygiene and infection control facilities, biomedical waste management and cleaning of the physical environment) ^3, 4^.

Sub-Saharan Africa is the region with the lowest levels of access to safe water and sanitation initiatives and the lowest rate of improvement in sanitation, with an estimated 695 million people still using the unimproved infrastructure. More specifically, in a research published in 2018, authors compiled data for 21 indicators of environmental conditions and standard precaution items from 78 low- and middle-income countries (LMICs), representing 129,557 HCFs. Results showed that 50% of HCFs lack piped water, 33% lack improved sanitation, 39% lack handwashing soap, 39% lack adequate infectious waste disposal, 73% lack sterilisation equipment, and 59% lack reliable energy services ^1^.

Mali, a continental country in West Africa, has minimal improvement in WASH. Indeed, in Mali, 50% of facilities do not have sufficient water storage and over 70% have water of poor quality. In addition, there is no national policies and plans on WASH in health care facilities ^5^.

In 2015, an intervention aiming to improve maternal and children environment in healthcare facilities (BECEYA) was launched in three regions of Mali. For five years, the project was a collaboration between the Government of Canada and the Ministry of Health and Social Affairs of Mali. BECEYA intervention components include the management of the hygiene and biomedical waste; the improvement of selected health facilities infrastructures mainly through four essential services: water supply (access points to drinking water, washing areas, shower), installation/rehabilitation of adapted latrines, solar energy electrification. As part of these essential components, the intervention aimed to reinforce the medical staff’s capacity to play their role in biomedical waste management through support to the national and regional health and public hygiene directorates. In addition, the implementation of this project involved the participation of various community actors such as community health and women’s associations.

This study aimed to explore the perceptions and experiences of patients and their companions, community actors and healthcare facilities staff on the effects of the BECEYA intervention in two regions of Mali.

## Methods

### Study design and setting

We conducted a qualitative study through individual semi-structured interviews and focus groups. Qualitative studies are indicated in research on social phenomena that are difficult to quantify ^6^. Therefore, an empirical phenomenological approach was used to obtain detailed descriptions about perceptions and experiences of patients and their companions, community actors and healthcare facilities staff on the effects of the BECEYA intervention. Phenomenological research describes commonalities of participants’ experiences across a population ^7^. The study was conducted in two Malian regions supported by the BECEYA project. As a result, two health centres were selected (Babala in Kayes and Wayerma 2 in Sikasso).

### Study participants and recruitment

Participants were recruited through purposive sampling. Variation in gender, age, occupation and function in the community or the health facility were considered to obtain diversity in the perceptions and experiences of the effects of the BECEYA intervention. The sample size was determined by data saturation (the point where no new themes from participants’ experiences emerged). These participants consisted of:

- women who attended antenatal care and maternity in the selected healthcare centres and their companions.
- health facility staff members (physician, midwives, nurses, care assistant, maintenance worker, hygiene and sanitation agent).

The healthcare staff members were recruited through each BECEYA project focal point. A designated nurse coordinator approached women attending antenatal care and their companions to request their participation in the study.

### Data collection

Data were collected during January and February 2020 through semi-structured individual interviews and focus groups.

Focus group interviews were conducted with the health centres’ staff concerned. In addition, semi-structured, individual interviews were conducted with women attending antenatal care and their companions, the manager of the healthcare centre.

The individual semi-structured interviews and focus groups guides were developed based on Donabedian’s conceptual framework adapted to the context of the BECEYA project (Donabedian, 1988). Interview guides were translated into the local language (Bambara) and tested with a population sample. With participant permission, all interviews were audio-recorded. Sociodemographic characteristics were collected at the start of the interview, followed by questions about participants’ experiences and perceived effects of the BECEYA project.

### Data analysis

The audio recordings were transcribed verbatim, translated into French, and reviewed by the interviewers for accuracy. All authors agreed and chose the highlighted quotations during the data analysis. All quotations were translated into English by PN. The analyses were carried out using the QDA Miner qualitative analysis software from Provalis. A thematic analysis was conducted in five main steps: 1) reading and familiarising with the data, 2) developing initial codes (coding), 3) searching for themes, 4) reviewing potential themes and 5) identifying categories of themes using a mixed (inductive and deductive) approach ^8, 9^. Two research team members (KR and MV) carried each of the steps in this process. We followed the Consolidated criteria for reporting qualitative research (COREQ) ^10^ throughout this study.

### Ethical considerations

Ethics approval for this study was obtained from the Institutional Ethics Committee of the Faculty of Medicine, Odontostomatology and Pharmacy of the University of Sciences, Techniques and Technologies of Bamako, Mali (N° 2020/06/CE/FMOS/FAPH). An information sheet explaining the research objectives was given and explained to each participant. Signed consent was required and obtained before each interview. Confidentiality was assured by using numbers instead of names and removing identifying information from the transcripts. All audio recordings and transcripts were saved on a password-protected computer.

## Results

Our sample consisted of 26 participants in individual interviews (20 women attending prenatal care and maternity services, 10 per health centre, four companions and two healthcare centre managers) and 21 participants in focus groups (21 healthcare centre staff members, 10 in Babala, 11 in Wayerma 2). Characteristics of women attending prenatal care and maternity services, companions, and healthcare staff are available in Tables 1, 2 and 3. Donabedian’s conceptual framework of quality of care ^11^ was used to present the perceived changes following the implementation of the BECEYA project. The themes are related to the infrastructure (perceived changes in the characteristics of the healthcare facilities setting, including the infrastructure introduced by the BECEYA project), process (changes in the delivery and use of care introduced or resulting from BECEYA activities) and outcome (the direct and indirect effects of these changes on the health status of patients and the population).

**Table 1.** Characteristics of women attending prenatal care and maternity services (n = 20)

|  | BABALA | W2 |  | BABALA | W2 |
| --- | --- | --- | --- | --- | --- |
| AGE (YEARS) |  |  | OCCUPATION |  |  |
| 15 - 20 | 4 | 2 | Housewife | 4 | 5 |
| 21 - 25 | - | 3 | Trader | 1 | 2 |
| 26 - 30 | 6 | 3 | Teacher | - | 1 |
| 31 - 35 | - | 1 | Dressmaker | 1 | 1 |
| 36 - 40 | - | 1 | Nurse | - | 1 |
|  |  |  | Farmer | 3 | - |
|  |  |  | Student | 1 | - |
| Total | 10 | 10 | Total | 10 | 10 |
| PLACE OF RESIDENCE |  |  | PLACE OF DELIVERY |  |  |
| Babala | 9 | - | Babala health centre | 8 | - |
| Wayerma 2 | - | 6 | Wayerma 2 health centre | - | 10 |
| Other | 1 | 4 | Other health centre | 2 | - |
| Total | 10 | 10 | Total | 10 | 10 |
| MATRIMONIAL STATUS |  |  | NUMBER OF PRENATAL VISITS |  |  |
| Married | 10 | 9 | 4 | 10 | 9 |
| Single | - | 1 | 5 |  | 1 |
| Total | 10 | 10 | Total | 10 | 10 |
| LEVEL OF EDUCATION |  |  | BABALA | WAYERMA 2 |  |
| None |  |  | 5 | 3 |  |
| Primary school |  |  | 4 | 3 |  |
| Secondary school |  |  | - | 2 |  |
| University |  |  | - | 2 |  |
| TOTAL |  |  | 10 | 10 |  |

**Table 2.** Characteristics of companions.

|  | BABALA | W2 |  | BABALA | W2 |
| --- | --- | --- | --- | --- | --- |
| AGE (YEARS) |  |  | SEX |  |  |
| 15 - 20 | 1 | - | Women | 1 | 1 |
| 21 - 25 | - | 1 | Men | 1 | 1 |
| 26 - 30 | - | 1 |  |  |  |
| 31 - 35 | - |  |  |  |  |
| 36 - 40 | - |  |  |  |  |
| 41 - 45 | 1 |  |  |  |  |
| Total | 2 | 2 | Total | 2 | 2 |
| PLACE OF RESIDENCE |  |  | OCCUPATION |  |  |
| Babala | 1 | - | Trader | - | 1 |
| Wayerma 2 | - | 1 | Farmer | 1 | - |
| Other | 1 | 1 | Bricklayer | - | 1 |
|  |  |  | Mechanic | 1 | - |
| Total | 2 | 2 | Total | 2 | 2 |
| MATRIMONIAL STATUS |  |  | FAMILY RELATIONSHIP |  |  |
| Married | 1 | 1 | Aunt | 1 | - |
| Single | 1 | 1 | Brother-in-law | 1 | 1 |
|  |  |  | Mother-in-law | - | 1 |
| Total | 2 | 2 | Total | 2 | 2 |

**Table 3.** Characteristics of healthcare staff.

| ÂGE (YEARS) |  |  | OCCUPATION |  |  |
| --- | --- | --- | --- | --- | --- |
| 20 - 25 | 1 | - | Matrons | 3 | 1 |
| 26 - 30 | 2 | 1 | Security guard | 1 | 1 |
| 31 - 35 | 1 | 5 | Manager | 1 | - |
| 36 - 40 | 2 | - | Assistant nurse | 1 | - |
| 41 - 45 | - | 1 | Cleaning agent | - | 1 |
| 46 - 50 | 2 | 3 | Nurses | - | 6 |
| 51 - 55 | 1 | - | Technologist | 1 | - |
| 56 - 60 | - | 1 | Midwife | 2 | 1 |
| 61 - 65 | 1 | - | Physicians | 1 | 1 |
| Total | 10 | 11 | Total | 10 | 11 |
| PLACE OF RESIDENCE |  |  | NUMBER O YEARS ON THE JOB |  |  |
| Babala | 8 | - | 0 - 5 | 3 | 6 |
| Wayerma 2 | - | 5 | 6 - 10 | 3 | 1 |
| Autre | 2 | 6 | 11 - 15 | 2 | - |
|  |  |  | 16 - 20 | 1 | 4 |
|  |  |  | 21 - 25 | 1 | - |
| Total | 10 | 11 | Total | 10 | 11 |
| MATRIMONIAL STATUS |  |  | SEX |  |  |
| Married | 10 | 11 | Men | 5 | 2 |
|  |  |  | Women | 5 | 9 |
| Total | 10 | 11 | Total | 10 | 11 |

### Perceived changes in infrastructure from the perspective of women attending antenatal care and their companions

Healthcare centres that benefited from the BECEYA project experienced many infrastructure changes. When comparing the physical structure before and after the BECEYA project, women attending antenatal care and their companions described their observed changes. It was difficult for one participant to describe all the existing changes fully, but the discourses were complementary.

The various changes in the infrastructure of the CSCom reported by the participants concern all the components of WASH, namely hygiene (provision of gender-specific latrines, new handwashing equipment i.e. devices consisting of a bucket of drinking water above a wastewater collection container and a holder for soap, the addition of a washing area for clothes, medical instruments, dishes, etc.); water (new water supply facilities including a water tower and drinking water taps); sanitation (the installation of the incinerator and the equipment for the management of biomedical waste, including the three-colour bins). In addition, electricity has been installed in certain sections of the health centre. The women users and their companions commented on their feelings about the changes brought about by these new infrastructures, particularly about the cleanliness of the health centres: “Before there was no maintenance in this centre, but today there is much hygiene in this centre, the latrines are cleaner too, every time I go into the latrines I see that they are clean” (W2_Wo9) or again: Before there was only one latrine for the whole centre, now it is the opposite, there are latrines for men, there are also some for women (B_Ac1). The changes in the infrastructure of the CSCom brought about by the BECEYA project have enabled women to give birth and professionals to work in a better-equipped environment. One user reported that “before there was no lighting in the centre, we delivered by the light of small torches” (B_ Wo9). A participant made the following remark on water availability in the health centres: “Before, water was not available in this centre. But now water is available in the centre’ (B_ Wo2)

It seems impossible for the women users and their companions to distinguish between the achievements of the BECEYA project and those of other projects. Indeed, the participants were asked about the changes in general that they had seen in their CSCom in recent years, not just the changes brought about by BECEYA. In participants’ interviews, we observed changes made simultaneously as the BECEYA project does not stem from it. For example, the improvement of the maternity material (new insecticide-impregnated mosquito nets in Wayerma 2 and plastic on the beds in Babala) and the increase of the number of beds in the rooms. These changes are not directly related to BECEYA’s activities but are cited by participants. According to them, they have observed these changes in their CSCom over the past few years.

Babala users reported that “before, women used to deliver on the table without spreading the plastic, but now he spreads the plastic on the table before delivering” (B_ Wo10), while in Wayerma 2, it is that “now new deliveries lie under clean mosquito nets. Otherwise, there was no net before. So, women were only lying on beds without nets. The distribution of insecticide-treated mosquito nets in the health centre is part of a major effort by the Ministry of Health to protect pregnant women and children aged 0-5 years from mosquito bites that could be dangerous for them.

Women attending ANC and their companions were asked about their perceptions of the new infrastructure. The participants expressed themselves according to the changes they appreciated the most. Some had a specific appreciation for a change. For example, for the availability of running water, one user reported: “…but what I appreciated most was the availability of water at the centre. Before, there was a water problem at the centre; when you came to give birth at the health centre, you couldn’t find water to wash yourself or your clothes. You can easily find water at the centre (B_ Wo1). Another participant rather appreciates the availability of electricity: “I appreciate electricity the most” (B_Ac1). Other participants have a more general appreciation of the new infrastructure, with one user stating: “there is not a single thing in this centre that I don’t appreciate. I appreciate everything in this centre” (B_ Wo6).

### Perceived changes in infrastructure from the perspective of health centre staff

The health centre’s staff also reported their perceptions of the various changes following the BECEYA project’s implementation. Their perceptions corroborate those of women attending antenatal care and their companions. They reported that new infrastructures implemented by the BECEYA project in the health facilities improved the quality of services and care. For example, one participant reveals that “Before the BECEYA project, there was only one latrine for the whole health centre. Thanks to BECEYA, the health centre staff, patients, and even the disabled have their latrines (one for men and one for women)” (B_FG_HCP).

Another participant spoke about the cleanliness of the working environment and the establishment of washing areas by the project; he said: “…the cleanliness of the centre, in general, whether the dispensary, the maternity ward, the incineration, the management of biomedical waste, the placentas, the washing areas, once the women have given birth, they wash, do the laundry, the soiled clothes, that is it. In general, it brings cleanliness to the centre (W2_HCM). Another participant gives his appreciation of the hygiene and cleanliness of the health centre: “Before the arrival of BECEYA, we put the waste in any way. Now, for every piece of rubbish lying around in the centre’s courtyard, we pick it up and put it in the appropriate container. We teach the women how to deal with waste. Instead of just throwing it in the yard, we show them the appropriate container for it” (B_ FG_HCP).

### Perceived changes in the care process from the perspective of women attending antenatal care and their companions’

The new infrastructure has led to changes in practices in the health facility. For example, women attending antenatal care and their companions’ were asked about the changes they had observed in their health centre regarding patient care. They described a cascade of changes in service users’ care practices and behaviours, women in their communities and staff. These changes result from the improved infrastructure at the two health centres.

They reported the excellent reception and quality of care received and the perceived competence of the staff in the health centre. These elements positively influence women’s experience at the health centre and encourage them to use the services, as reported by one user: “I appreciate the reception, when I came straight away a midwife welcomed me well, she took my papers and guided me to the consultation room, and I liked that a lot” (W2_Wo10). Another participant spoke of the quality of care, particularly the post- partum follow-up, which she said had improved: “Nowadays when you give birth in the centre, you stay under surveillance for six hours before leaving for home, which we didn’t do before. Before BECEYA, some women left for home right after their delivery” (W2_Wo7).

One woman reports the role of the incinerator in the management of the placenta after delivery as follows: “The construction of the incinerator has helped us a lot in the management of the placenta, often when you give birth it was tough to find the old women to bury it. When you give birth in the morning, you could spend the whole day with the placenta without finding a person to bury it. However, this difficulty is over thanks to the incinerator” (W2_Wo8).

Finally, participants reported changes in personal hygiene practices such as handwashing due to the new infrastructure put in place by the BECEYA project. Indeed, women attending antenatal care and their companions’ reported the importance of implementing handwashing recommendations by all. For one participant, “the first change I noticed was the application of handwashing with soap in the centre” (B_Wo1).

### Process changes from the perspective of health centre staff

The changes observed and reported by the health staff affect access to care for the poorest and the supply of essential medicines and inputs. For example, the reduction in delivery costs is reported by one participant: “delivery costs were prohibitive here, but this is not the case now” (B_ FG_HCP). In addition, health centres are better supplied with inputs to provide care to clients. This motivates clients to visit the health centre more often, as staff members report in the following excerpts: “Mothers accompany their children to the ComHC when they are ill. Medicines are available at the health centre (W2_Personnel 3).

“Women no longer take sick children home with them. As soon as children fall ill, they are taken to the health centre” (W2_ FG_HCP).

Participants also perceived changes in the women’s involvement in sanitation at the health centre. To illustrate this, one participant reported: “…they have changed, because every Saturday they come here to the centre to wash our rooms, sweep the courtyard, and then every 15th of the month it is the village sanitation day…” (B_HCM).

### Perceived effects on health outcomes from the perspective of women attending antenatal care and their companions’

The majority of participants perceived the positive impacts of this project on the health of women and children. The project has raised awareness among women users. As a result, they are motivated to use health services, especially in the health centre supported by BECEYA. The changes observed have led to increased awareness of the importance of attending health centres during pregnancy and favouring assisted childbirth. According to one respondent, “the changes I have observed are that ANC is beneficial for women. Moreover, it makes childbirth easier” (B_Ac2).

Another participant reports that “before, women preferred to give birth at home rather than coming to the centre. But now they come to give birth at the centre” (B_Ac1).

The awareness and participation of women in health-related activities have led to changes in the health of community members. In this regard, one user reported the following about hygiene awareness and new care practices: “The application of advice provided by health workers has reduced several diseases, the application of bleach has also reduced diarrhoeal diseases” (B_Wo). Another user reports the following on the impact of environmental sanitation: “these changes have influenced a lot, you know, many diseases are related to mosquitoes and sewage; if these things decrease the disease too, decreases. So these changes have decreased many diseases in the community” (W2_Wo).

### Perceived effects on health outcomes from the perspective of the health centre staff

The health centres that benefited from the BECEYA project saw their activities increase, mainly thanks to the awareness-raising done in the community. As an illustration, one participant reports: “the sensitisation has changed many things in the work, especially in prenatal consultations, vaccinations, deliveries, family planning and postnatal consultations. With the arrival of BECEYA, these activities have expanded” (B_ FG_HCP). Another participant added: “After the sensitisation, we have noticed that women are increasingly vaccinating their children. In addition, women, after having known the advantages of family planning through sensitisation, no longer hesitate to use family planning” (W2_ FG_HCP).

The BECEYA project has also had positive results in terms of gender relations. Men have realised the need to give their wives more freedom to enjoy by being involved in awareness-raising activities. One participant reports that ‘there has been a change, as before men did not accept their wives to advocate or speak in public. However, this has changed since the arrival of BECEYA. The proof is that today the women of Babala have gone to another village to raise awareness, even some men accompany them because they have understood that it is a good thing” (B_ FG_HCP).

## Discussion

This study aimed to analyse the perceptions and experiences of service users and health staff on the effects of a project to improve the health environment of community health centres in two regions of Mali.

The study results provided an overall picture of the changes observed in the health centres supported by the BECEYA project, from the perspective of the women users of the services, their companions and the health centre staff. One of the changes repeatedly reported in both health centres was the environment’s cleanliness and salubrity. In terms of infrastructure, the vast majority of participants at both health centres noted the same changes, i.e. the addition of gender-specific latrines, the addition of electricity in some sections of the centre, new water infrastructure, improved delivery room equipment, the addition of the incinerator and biomedical waste management, new handwashing equipment, the addition of a washing area, and the increased number of beds in the rooms. All of these were seen as positive and associated with an increase in the quality of services. In addition, most users noted the improved care practices and additional services.

Furthermore, the staff of the two health centres spoke of strengthening their work capacity, as they have more means at their disposal, such as medicines in the centre’s pharmacy, permanent electricity, and a clean environment. According to the staff, changes in infrastructures and healthcare professionals’ practices increased women’s antenatal care use. The latter emphasised the existence of an unprecedented motivation among women to use health services and their commitment to maintaining cleanliness in their living environment. Overall, changes in infrastructures and processes had a positive impact on the health of patients and the general population. Participants in all groups reported an increased awareness of their health and influenced the centre’s services. There was also an awareness of seeking care at the onset of symptoms and care related to pregnancy and institutional childbirth.

### Comparison with the literature

Barriers to the use of hospital care are mainly related to the quality of care, especially for maternity services (often inadequate, unaffordable, under-staffed and lacking in medically trained professionals) ^12^. Water, hygiene and sanitation (WASH) services in health facilities help respond to outbreaks and prevent healthcare-associated infections. They are essential to ensure healthcare quality ^5, 13^. In resource-limited countries, lack of access to affordable energy and adequate clean water in health facilities is a major factor in high maternal and infant morbidity and mortality ^3, 14-16^. Lack of electricity makes it impossible to operate cold chains capable of storing life-saving vaccines ^16^, while inadequate drinking water impacts sanitation, where infectious diseases can develop and spread ^3, 17^. This makes it difficult to access essential maternal and child health services timely and affordable ^14, 15, 18^.

Furthermore, inadequate WASH services in health care facilities have also been shown to be associated with increased patient dissatisfaction and even a barrier to service utilisation in some settings (including maternity services) ^3, 13, 18^. Indeed, the lack of high-quality WASH facilities in delivery rooms is often cited as to why women preferred home delivery ^3, 18^. Women expect health facilities to have an adequate WASH system, essential for their human rights, dignity, and infection prevention. However, achieving this goal remains a distant prospect in many health facilities in developing countries ^18^. Satisfaction with health services is defined as the extent to which patients positively perceive the care provided by nurses or medical staff ^19^. Patient satisfaction reveals the extent to which health care needs are being met and provides a key indicator of high-quality health care. In addition, it is a factor used for planning and evaluating health interventions ^19^.

Numerous studies have shown that patients who are satisfied with the care provided by health personnel are more likely to use health services in the future and to comply with the prescribed medical treatment until the end. Therefore, for patients to be more satisfied with the treatment, there is a need to provide high-quality healthcare considered safe, timely, effective, efficient, equitable and patient-centred ^18, 19^.

Providing high-quality care in maternity services involves giving mothers the best possible medical care and outcomes during the antenatal, delivery and postnatal period that can be measured against standard guidelines ^20^. Evidence shows that pregnant women are more likely to give birth in health facilities if satisfied with their care at service delivery points ^21-23^. However, other studies have shown that poor quality of services and negative attitudes of health workers prevent pregnant women from using these services ^21, 24^. Therefore, in addition to improving facility infrastructure, quality of care, and cost-effectiveness, improvements in maternity services should also address providers’ interpersonal attitudes and behaviours ^24, 25^.

### Importance for research and practice

It was relevant to have data on the perceptions and experiences of the key stakeholders to demonstrate that the effects of the BECEYA project go beyond improving the health environment. The results also show that the BECEYA project has brought about profound changes in the health facilities. These changes have positively influenced the motivation to attend the health centres by women service users and improve the quality of care. However, despite the significant effects of the BECEYA project, structural barriers prevent all women from benefiting from the best pregnancy care. One of these major barriers is the lack of financial resources to meet the cost of the services ^25^. Indeed, the implementation of strategies designed to encourage attendance at health centres for antenatal care worldwide tends to be based on the assumption that people will attend if there is a good quality of services. However, sociodemographic data indicate that women from relatively poor backgrounds, living in rural areas and/or with low levels of education are less likely to access antenatal services, even if they are available ^26^. In addition, the maintenance and upkeep of infrastructures in place and the ongoing staff training also need to be addressed as they ensure the sustainability of these interventions ^3^.

### Limitations

The choice of the two health centres studied was not made randomly, and the perceptions analysed only reflect the realities of these two contexts. Furthermore, to please, the participants may have responded according to social desirability.

## Conclusion

This study aimed to evaluate the effects of a project to improve the health environment for mothers and children in health centres. The study identified the positive effects of women users of the services, their companions and health centre staff following the implementation of the project. The changes brought about by the project are perceived both in terms of infrastructure (WASH services, electricity, cleanliness) and processes (reception, availability of staff and working materials, inputs and medicines). These changes have had positive results for women attending antenatal care and their companions’ (better care experience, improved quality of care), for health centre staff (improved environment and working conditions). Therefore, this study contributes to the advancement of knowledge to show the link between improving the health environment of health centres in developing countries, a major aspect of the quality of care, and maternal and child health care.

## Data Availability

All relevant data are within the manuscript and its Supporting Information files

## Acknowledgements

We would like to thank all the participants for taking part in the study. We would also like to thank Dr Aissatou Tinka Bah, Ericka Moerkerken for their support in carrying out this study and the Sante Monde (CCISD) team in Mali for their logistical support during data collection.

## Funding

This study was funded by Sante Monde (CCISD) (https://santemonde.org/) through Global Affairs Canada. The funding body was not involved in the study design, collection of data, analysis and interpretation of data, writing of the manuscript or the decision to submit the manuscript for publication.

## Competing interests

None of the authors have any financial or conflict of interest to declare

## Authors’ contributions

Authors contributions include the following: Sante Monde - CCISD (funders) conceived the original idea for the study. PN, KR and MV developed the research protocol. PN was responsible for the study design. BAL offered guidance on qualitative methodology and revising the manuscript. FT was responsible for interviewing participants. KR and MV carried out the qualitative analysis of interview data. All the authors have read and approved the final manuscript.

## Notes

### Competing Interest Statement

The authors have declared no competing interest.

### Funding Statement

No: The funders had no role in study design, data collection and analysis, decision to publish, or preparation of the manuscript.

## References

1. Cronk R and Bartram J. Environmental conditions in health care facilities in low- and middle-income countries: Coverage and inequalities. International Journal of Hygiene and Environmental Health 2018; 221: 409–422. DOI: 10.1016/j.ijheh.2018.01.004.

2. United Nations. Transforming our world: the 2030 Agenda for Sustainable Development. 2015. New York.

3. World Health Organization and the United Nations Children’s Fund. WASH in healthcare facilities - Global Baseline Report 2019. 2019. Geneva.

4. Cronk R, Guo A, Folz C, et al. Environmental conditions in maternity wards: Evidence from rural healthcare facilities in 14 low- and middle-income countries. International Journal of Hygiene and Environmental Health 2021; 232: 113681. DOI: 10.1016/j.ijheh.2020.113681.

5. World Health Organization. Water, sanitation and hygiene in health care facilities - Status in low- and middle-income countries and way forward. 2015. Geneva.

6. Hammarberg K, Kirkman M and De Lacey S. Qualitative research methods: when to use them and how to judge them. Human Reproduction 2016; 31: 498–501. DOI: 10.1093/humrep/dev334.

7. Groenewald T. A Phenomenological Research Design Illustrated. International Journal of Qualitative Methods 2004; 3: 42–55. DOI: 10.1177/160940690400300104.

8. Bradley EH, Curry LA and Devers KJ. Qualitative Data Analysis for Health Services Research: Developing Taxonomy, Themes, and Theory. Health Services Research 2007; 42: 1758–1772. DOI: 10.1111/j.1475-6773.2006.00684.x.

9. Braun V and Clarke V. Thematic analysis. In: H. Cooper PMC, D. L. Long, A. T. Panter, D. Rindskopf, & K. J. Sher (ed) APA handbook of research methods in psychology, Vol 2 Research designs: Quantitative, qualitative, neuropsychological, and biological American Psychological Association, 2012, pp.57–71.

10. Tong A, Sainsbury P and Craig J. Consolidated criteria for reporting qualitative research (COREQ): a 32-item checklist for interviews and focus groups. International Journal for Quality in Health Care 2007; 19: 349–357. DOI: 10.1093/intqhc/mzm042.

11. Donabedian A. The quality of care. How can it be assessed? JAMA 1988; 260: 1743–1748. 1988/09/23. DOI: 10.1001/jama.260.12.1743.

12. Izugbara CO and Wekesah F. What does quality maternity care mean in a context of medical pluralism? Perspectives of women in Nigeria. Health Policy and Planning 2018; 33: 1–8. DOI: 10.1093/heapol/czx131.

13. Weber N, Martinsen AL, Sani A, et al. Strengthening Healthcare Facilities Through Water, Sanitation, and Hygiene (WASH) Improvements: A Pilot Evaluation of “WASH FIT” in Togo. Health Security 2018; 16: S-54-S-65. DOI: 10.1089/hs.2018.0042.

14. Dalinjong PA, Wang AY and Homer CSE. Are health facilities well equipped to provide basic quality childbirth services under the free maternal health policy? Findings from rural Northern Ghana. BMC Health Services Research 2018; 18. DOI: 10.1186/s12913-018-3787-1.

15. Essendi H, Johnson FA, Madise N, et al. Infrastructural challenges to better health in maternity facilities in rural Kenya: community and healthworker perceptions. Reproductive Health 2015; 12. DOI: 10.1186/s12978-015-0078-8.

16. Franco A, Shaker M, Kalubi D, et al. A review of sustainable energy access and technologies for healthcare facilities in the Global South. Sustainable Energy Technologies and Assessments 2017; 22: 92–105. DOI: 10.1016/j.seta.2017.02.022.

17. Watson J, D’Mello-Guyett L, Flynn E, et al. Interventions to improve water supply and quality, sanitation and handwashing facilities in healthcare facilities, and their effect on healthcare-associated infections in low-income and middle-income countries: a systematic review and supplementary scopin. BMJ Global Health 2019; 4: e001632. DOI: 10.1136/bmjgh-2019-001632.

18. Bouzid M, Cumming O and Hunter PR. What is the impact of water sanitation and hygiene in healthcare facilities on care seeking behaviour and patient satisfaction? A systematic review of the evidence from low-income and middle-income countries. BMJ Global Health 2018; 3: e000648. DOI: 10.1136/bmjgh-2017-000648.

19. Asadi-Lari M, Tamburini M and Gray D. Patients’ needs, satisfaction, and health related quality of life: Towards a comprehensive model. Health and Quality of Life Outcomes 2004; 2: 32. DOI: 10.1186/1477-7525-2-32.

20. World Health Organization. Standards for improving quality of maternal and newborn care in health facilities. 2016. Geneva.

21. Chukwuma A, Wosu AC, Mbachu C, et al. Quality of antenatal care predicts retention in skilled birth attendance: a multilevel analysis of 28 African countries. BMC Pregnancy and Childbirth 2017; 17. DOI: 10.1186/s12884-017-1337-1.

22. Lothian JA. Safe, Healthy Birth: What Every Pregnant Woman Needs to Know. Journal of Perinatal Education 2009; 18: 48–54. DOI: 10.1624/105812409x461225.

23. Nyongesa C, Xu X, Hall JJ, et al. Factors influencing choice of skilled birth attendance at ANC: evidence from the Kenya demographic health survey. BMC Pregnancy and Childbirth 2018; 18. DOI: 10.1186/s12884-018-1727-z.

24. Mannava P, Durrant K, Fisher J, et al. Attitudes and behaviours of maternal health care providers in interactions with clients: a systematic review. Globalization and Health 2015; 11. DOI: 10.1186/s12992-015-0117-9.

25. Oyugi B, Kendall S and Peckham S. Effects of free maternal policies on quality and cost of care and outcomes: an integrative review. Primary Health Care Research & Development 2021; 22. DOI: 10.1017/s1463423621000529.

26. Finlayson K and Downe S. Why Do Women Not Use Antenatal Services in Low-and Middle-Income Countries? A Meta-Synthesis of Qualitative Studies. PLoS Medicine 2013; 10: e1001373. DOI: 10.1371/journal.pmed.1001373.

